# Causal interhemispheric neuromodulation sharpens synaptic and neurobehavioral inhibition in stroke

**DOI:** 10.1101/2025.03.10.25323712

**Authors:** Joao Castelhano, Felix Ducker, Carolina Xavier, Nádia Canário, Isabel Duarte, Sónia Afonso, Ana Gabriel Marques, Cecilia Lourenço, Filipe Carvalho, Jorge Lains, Angela Neves, João Sargento Freitas, Gustavo Santo, Antero Abrunhosa, Alexander T. Sack, Miguel Castelo-Branco

## Abstract

Hemispheric functional asymmetries are a key aspect of human brain organization. Revealing how causal manipulations of hemispheric (im)balances dynamically change pre and postsynaptic GABA regulation, in terms of mean and variance changes, is essential to understand plasticity of brain asymmetries in health and disease. This has direct implications for treatment approaches in stroke relying on interhemispheric inhibition using transcranial magnetic stimulation (TMS). We investigated neurobehavioral and neurochemical effects of cortical stimulation using a unique multimodal brain imaging and stimulation set-up, in both healthy and unilaterally lesioned hemispheres, in stroke. We performed two molecular imaging sessions measuring GABA receptor levels, with sham and real neurostimulation, fMRI and GABA neurospectroscopy, all in the same day. We found that noninvasive inhibitory neuromodulation of interhemispheric interactions causes pre-postsynaptic sharpening of GABA neurotransmitter and receptor levels, providing a mechanism of action for TMS contralateral inhibitory protocols in stroke. This discovered plasticity mechanism based on correlated reduction of pre and postsynaptic variability represents a basic principle underlying long range inhibitory hemispheric interactions and allows for stabilization of synapses and re-balancing of brain activity in disorders of hemispheric imbalance, such as stroke.

## Introduction

Functional differences between the left and right hemisphere are present in many species and there are countless examples of behavioral manifestations of this lateralization principle of brain organization^1^ ^2^. Although hemispheric lateralization has been established across multiple cognitive domains, the underlying nature of hemispheric interactions and resulting asymmetries remain poorly understood. In particular, the relation to synaptic mechanisms are unknown and a reduction to basic biological principles is therefore lacking. The biological principles underlying these hemispheric asymmetries and their relation to synaptic mechanisms, however, remain unknown. Unravelling how interhemispheric organizational principles are related to molecular regulation of synaptic function is instrumental in understanding one of the most basic principles of human brain organization as well as functional deficits related to acquired hemispheric imbalances.

Transcranial magnetic stimulation (TMS) can be used to experimentally modulate hemispheric interactions and induce lasting neuroplastic changes in hemispheric asymmetries. However, while established on behavioural and neural network levels, we still lack direct evidence by which exact biological and molecular mechanisms these effects are caused and hence by which neurobiological principles these general hemispheric functional asymmetries are organized ^3^.

Cellular and computational studies have shown that synaptic parameters can be derived not solely from mean values but that actually, variance, and covariance in neural measures are a rich source of information^4^. Accordingly, variance analysis can be used as a tool to predict the mechanism underlying synaptic plasticity^5^. Importantly, modelling work has suggested that variability of postsynaptic responses depends non-linearly on the number of synaptic inputs ^6^, which may be changed by transcranial stimulation. Whether these principles apply to humans remains an open question. A recent study suggested that this might be the case, by showing that increased variability of excitatory drive to fast-spiking parvalbumin interneurons, as suggested by postmortem molecular studies in schizophrenia, might lead to lower prefrontal gamma power, as further suggested by a computational model ^7^.

In this line, we hypothesize that synaptic variability plays a critical role in hemispheric asymmetries where postsynaptic response optimization at inhibitory synapses might lead to the desired optimal excitation/inhibition balance between hemispheres through variance reduction, in line with prior modelling and cellular work ^8^. Long-term modifications of neuronal connections are critical for learning and plasticity. Their locus of expression - pre- or postsynaptic - and respective covariance remains debated. In line with the above-mentioned theoretical framework short and long-term plasticity may depend on optimization of the postsynaptic response statistics toward a given mean with minimal variance. The state of the synapse may depend on the ratio of pre- and postsynaptic modifications. Such changes in pre and post synaptic variability may also be the driving force underlying various forms of neuroplasticity ^9^. Approaches to re-balance the hemispheres to alleviate functional deficits resulting from dysfunctional hemispheric interactions provide a way to infer on biological significance. Conceptual models and combination of causal approaches, based on neurostimulation and/or lesion studies are instrumental in this respect. Combining TMS with unilateral lesion studies (e.g. unilateral stroke models) provides a unique opportunity to test specific theories of interhemispheric interaction ^10,11,12^ and their molecular basis. Our previous studies in the motor cortex of stroke patients show that cTBS can be used as an inhibitory protocol for the unaffected hemisphere^13–15^. We also have fMRI evidence for the interhemispheric inhibition model^16,17^. This has direct implications for treatment approaches stroke relying on interhemispheric inhibition using transcranial magnetic stimulation (TMS).

The interhemispheric inhibition model postulates that both hemispheres are directly interconnected and mutually controlling in a domain specific manner, i.e. inhibiting each other within an equilibrium of an optimally tuned excitation/inhibition balance. After unilateral brain damage, the competition between hemispheres is highly biased leading to decreased activity in the lesioned hemisphere but also increased activity in the contralesional hemisphere due to disinhibition ^18,19^. The result is an imbalance between hemispheres with large functional consequences ^11^. In this context, lateralized deficits are not merely due to a loss of function of the lesioned hemisphere but may also be the result of disrupted interactions between hemispheres at a synaptic level, further amplifying and consolidating these deficits. This provides a perfect model to experimentally study dynamic processes of hemispheric lateralization and functional asymmetries and how the changes in synaptic variability can be implicated in the process.

TMS has been proposed as a technique that can be used to selectively modify brain hemispheric (im)balance ^10,11^. It provides a noninvasive, reversible, and relatively localized approach that has substantial promise for particular clinical applications ^20,21^ including stroke rehabilitation^22^. However, the effects of TMS and the link between molecular, neurochemical, physiological and behavioral responses ^18,19^ are still enigmatic, in particular in the context of inter-hemispheric interactions. Here we uniquely combined multimodal and dual causal (TMS/lesion) strategies to address these questions. Unilateral stroke is a condition which provides a unique opportunity to test the hemispheric (im)balance hypothesis because it allows for the direct assessment of the interaction between a healthy and lesioned hemisphere within the same participant. Such combination of approaches allowing for deep physiological dissection is hard to achieve in healthy individuals ^14^. The current approach aims at understanding the underlying imbalance at a molecular level, following the above mentioned synaptic plasticity hypothesis, which is pivotal to understand principles of human brain organization underlying lateralization and to implement novel model-based treatment approaches, in disorders such as stroke where treatment protocols based on interhemispheric inhibition have been proposed ^23^.

## Materials and methods

### Study Experimental Design

We combined transcranial magnetic stimulation (TMS), magnetic resonance imaging (MRI) and positron emission tomography (PET) to experimentally manipulate the (im)balance between hemispheres. The experiment included a full day of testing with the following acquisitions (supplementary Fig. S1 Timeline): patients arrived from the rehabilitation Hospital to our institute in the morning and started a full neuropsychological evaluation, followed by a medical interview to evaluate the conditions to proceed with the exams. Next, they were prepared for PET imaging and performed a short assessment of resting motor threshold (rMT) with TMS. Then, a first TMS (real or sham) was applied followed by a PET- 11C- Flumazenil imaging acquisition and one MRI acquisition (including anatomical, functional and Spectroscopy recordings). After MRI, patients rest for about one hour and a second TMS (real or sham) was applied immediately before the second PET imaging. Although this was an extensive protocol all patients were very collaborative and able to perform the experiment. Further technical details on the procedures are described in the subsections below.

### Participants

Patients inclusion criteria were based on the following scores from the National Institutes of Health Stroke Scale: presence of extinction and inattention (item 11 > 0); first screening for the presence of neglect; absence of hemianopia (item 3 = 0, unless in case of extinction) to exclude perceptual deficits; absence of severe aphasia (item 9 <= 1) to ensure sufficient communication abilities; Sufficient strength in right arm (item 5 <= 1) to ensure that patients can do tasks with their right hand.

Nineteen stroke patients (9 females; mean age = 66.71 years; supplementary table S1), right-handed and with an isquemic lesion in the right hemisphere of the brain, were informed of the objectives and potential risks of the study and signed a written consent inform. The study was approved by the Ethics Committee of the Faculty of Medicine of the University of Coimbra and was conducted in accordance with the Declaration of Helsinki.

### Neuropsychological Evaluation

Behavioral measures were acquired in a neuropsychological evaluation comprising the LANDMARK TEST; TokenTest; Montreal Cognitive Assessment (MoCA); and Behavioral Inattention Test.

### Brain Stimulation

Transcranial magnetic stimulation (TMS) was delivered using a MagPro X100 magnetic stimulator connected to a round coil (MCF-125; MagVenture, Denmark). We placed the coil at the hotspot over the primary motor cortex area, at 45° to the sagittal plane and identified individual resting motor thresholds (rMTs). Then, we target the contralesional hemisphere (left Intra-parietal sulcus (IPS)) with a stimulation protocol that is well-known to decrease cortical excitability.

The TMS protocol was single blinded and as follows: 1) Determine resting motor threshold over left primary motor cortex. An observable muscle twitch in the right index finger or thumb was used as a criterion. First, we found the presumed hotspot with trial and error, varying both TMS coil position and TMS intensity. Then, we determined the resting motor threshold, defined as the lowest TMS intensity that produces a twitch in at least 5 out of 10 trials. 2) Determine stimulation site over parietal cortex: given the large experimental load for the participants, we relied on the international 10/20 electrode system to guide our coil positioning. Specifically, the round coil was positioned over P3 and oriented with the help of a template of the TMS coil at the correct location. To ensure successful brain stimulation in all our patients, we decided to establish a compromise by using a somewhat less focal round TMS coil, but ensuring more focality than most round coils. Traditional round coils have the strongest magnetic field at the out rim. However, we used a round coil that is wired in a way resulting in a magnetic field that is strongest at the center of the coil. 3) TMS protocol: a continuous theta burst protocol was applied, that is, 50Hz triplets at a 5Hz for 40 seconds (= 600 biphasic pulses; Fig. S1 B). This is an inhibitory protocol applied to the healthy hemisphere to test our hypothesis that reducing the activity of this hemisphere can increase the activation at the contralateral hemisphere (the right hemisphere in our case). The stimulation intensity was defined as 80% of the individual rMT (mean rMT=45.97 ± 3.09 %) that is associated with more consistent theta burst effects ^24^ and TMS studies in clinical populations tend to favor the use of the resting motor threshold when applying theta burst protocols, with actually less variable effects ^25,26^. Since the gap between TMS application and the PET scan should be as short as possible, the TMS protocol was performed while the subjects were already positioned in the PET scanner and initiated as late as possible, before the tracer is administered (9 min: 31s on average). During stimulation, the coil was maintained at the target point using an adjustable coil holder. TMS real (real stimulation) and TMS sham (sham stimulation) were applied before each PET scan in a counterbalanced order. The sham was applied with exactly the same protocol, with the coil close to patient’s head but oriented to the scanner bed instead of the participant head. This way we ensured a ‘feeling of being stimulated’ without stimulation for the sham recordings. The patients were not able to distinguish between real and sham conditions (self-report at the chance level).

### Image Acquisition

#### PET imaging

Patients receive TMS (sham and real stimulation; single blinded) each one followed by a CT scan and a 3-dimensional PET of the entire brain (90 slices, 2-mm slice sampling; Philips Gemini GXL). We used PET 11C-flumazenil (FMZ) imaging to measure GABA-A receptor binding in all stroke patients. Flumazenil is a competitive antagonist of the GABA-A receptor and it binds to the benzodiazepine site of the receptor. This binding site is largely and predominantly located in the postsynaptic compartment ^27^. Each dynamic list mode PET 11C-Flumazenil scan was acquired over a period of 60 minutes, after each TMS session.

#### MRI

All patients performed 3T MRI (anatomical scan, fMRI with a simple saccade paradigm and spectroscopy). Scanning was performed using a 3T Tim Trio (Siemens, Erlangen, Germany) with a 12-channel head coil for signal reception. T1-weighted structural images of the brain were acquired with a Magnetization Prepared Rapid Gradient Echo Imaging (MPRAGE) sequence with: 1 mm3 isotropic voxel, echo-time (TE)=3.42 ms; repetition time (TR)=2530 ms; slices=176; field-of-view (FoV)=256 mm; flip angle (FA)=7.0; inversion time 1100 ms; 256 × 256 matrix; and GRAPPA acceleration factor = 2. fMRI data was acquired with the following parameters: repetition time TR=2000 ms; echo time TE=30 ms; slices=37; flip angle FA=90°; field of view FoV=192 mm; slice thickness=3 mm; number of volumes=169. The fMRI run lasted 5 min 40 s.

Two H-MRS voxels were positioned one in the left IPS and the second in the right IPS (Fig. 3 A) We used the Hadamard Encoding and Reconstruction of MEGA-Edited Spectroscopy (HERMES) approach to quantify GABA and Glx. The sequence parameters were as follows: volume 30×30×30 mm; TR=2000 ms; TE=80 ms; Averages=392; FA=90; water suppression bandwidth 50 Hz; delta frequency=-1.7 ppm; edit pulse 1 frequency =1.9 ppm; edit pulse 2 frequency =4.56 ppm; edit off frequency=7.5 ppm. Two spectra with and without water suppression were recorded for further spectrum normalization.

#### fMRI task description

During functional acquisitions, the scanner room was dimmed and participants were asked to perform a saccade each time a target appear in the screen. During the baseline blocks (11 blocks; 8 TRs of duration each) subject had to fixate a cross in the center of the screen. In the saccade blocks (10 blocks, 8 TRs each) a white dot appear in the screen for 500 ms randomly presented at 8 distinct locations (2 up, 2 down, 2 left, 2 right) and subjects had to perform a saccade towards the target each time it appears (Fig. S1 C). Saccades were purely driven by the information provided by the target dot showing up thus strongly emphasizing voluntary control mechanisms, the voluntary execution of saccades. Stimuli were presented on a 40-inch LCD Nordic NeuroLab monitor at the back of the scanner, 1825 mm viewing distance. The video mode was 1920×1080 at 60 Hz, and background luminance was 100 cd/m2. The Presentation software package (NeuroBehavioural Systems, Albany, CA, United States) was used to control stimulus presentation. To confirm the compliance with the task, we recorded eye movement data (Eyelink 1000 plus, SRResearch, Canada) during the experiment at a sampling rate of 1 kHz.

### Image data processing and analysis

#### PET processing

PET data were reconstructed using a LOR-RAMLA algorithm. PET 11C-Flumazenil voxelwise binding potential (BP) was obtained using in-house made software (Fig. 1), with the modified reference tissue method 2 (MRTM2) as compartmental model and the pons as reference. PET and MRI data were normalized to Montreal Neurological Institute (MNI) space, the same geometric transformation was used, after the PET scan had been rigidly coregistered with the correspondent anatomical MRI scan. We performed whole-brain and region-of-interest (ROI)-based analyses. ROI-based quantification was performed in regions used for spectroscopy (Intra-parietal Sulcus - IPS - left and right).

**Figure. 1.**
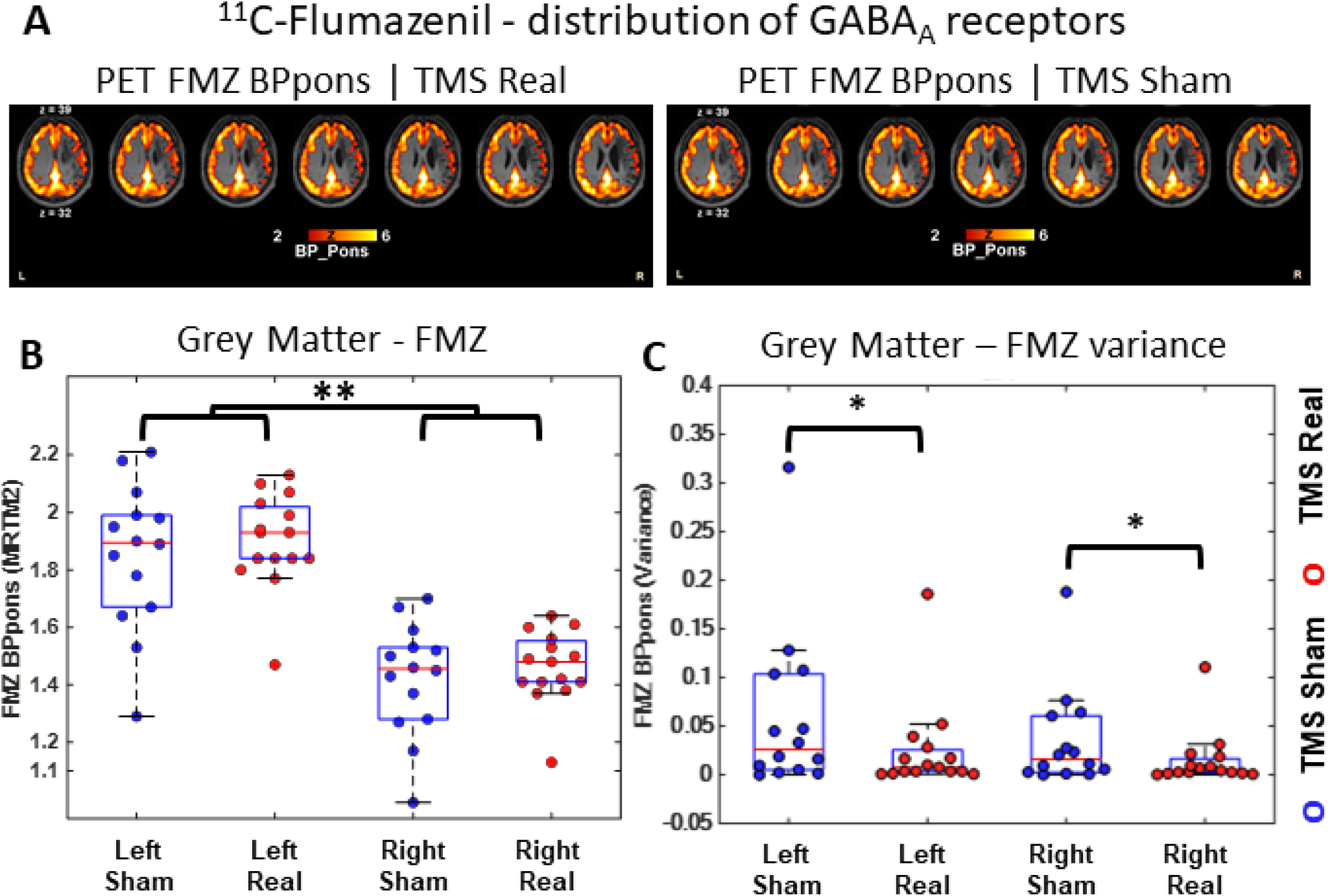
PET 11C-FMZ shows TMS induced global reduction of GABA receptor binding variance. (**A**) Quantification of PET data reveals the distribution of GABA_A_ receptors in the brain. A group average binding potential (BP) map is shown for TMS real and sham. (**B**) FMZ quantification in Whole brain Grey Matter (GM) revealed a significant reduction of GABAa receptors in the right (lesioned) hemisphere GM in comparison to healthy hemisphere (**P<0.0001) as expected due to the lesion and most importantly (**C**) a significant effect of the TMS in reducing the groups’ variance (Levene F(26)=4.62, * P=0.041, d=0.26).

#### MRS processing

Quantification of MRS data was performed using the Gannet 3.0 ^28^. GABA, Glx and Glutathione in addition to creatine signals were obtained from the difference edited spectra. The peaks for each metabolite were fitted to a simple Gaussian model. Creatine signal was fitted to a double Lorentzian model. All spectra had relative fit error <15%. GABA signal is known to be contaminated by other macromolecules ^29^. To account for differences in voxel tissue composition, the voxel fraction of White Mater (fWM) and grey mater GM (fGM) was used to normalize the GABA+ concentration according to: GABA+corr= GABA+/(fWM +fGM). An example output is shown in Figure 3.

#### Processing of structural images

We performed preprocessing of structural MRI data using BrainVoyager Qx (Brain Innovation) with the default pipeline settings. The brain representations in the figure panels were also created in this software. Structural images were normalized to AC-PC space. Raw image data is available from our institution repository upon request and supplementary Fig S2 show the group lesion region map.

#### Processing of functional images

The functional images were pre-processed with the default pipeline settings. 3D body motion correction, aligning all subsequent functional runs to the closest one to the anatomical scans, and temporal high-pass filtering (GLM-Fourier with two cycles sine/cosine per run, including linear trend removal) were applied to the functional data. Furthermore, the preprocessed images were registered to the structural space. Neural responses to the saccade task were assessed by applying a general linear model (GLM) analysis ^30^. Predictors for each condition were created by convolving the stimulation blocks with a standard hemodynamic response function ^31^. The fixation blocks were considered as baseline. Data were normalized with z-transformation and corrected for serial correlations with a second-order autoregressive method ^32^. A repeated measures ANOVA (factors: task and stimulation) was used to test for the effect of TMS on brain activation. A group GLM analysis was used to reveal the neural network associated to the saccades task for both TMS real and sham conditions.

### Statistical Analysis

Throughout the manuscript, and unless stated otherwise, significance for all fMRI results was assessed using cluster inference with a cluster-defining size of 300 voxels and a cluster probability of P < 0.05 FDR-corrected for multiple comparisons. Correlations were calculated with the Spearman correlation algorithm. The variances were calculated by taking the average of the squared differences of each data point from the sample’s mean for each parameter (e.g. PET: using each voxel of the ROIs; GABA measures variance were across subjects). The differences in the variances were tested with Levene’s test and comparison between conditions tested with Mann-Whitney at a significant threshold of 0.05 (FDR corrected for multiple comparisons).

### Data availability

All data and materials description are available in the main text or the supplementary materials. Furthermore, data, code, and materials used in the analysis are available upon request to the corresponding author.

## Results

We started by the quantification of the whole brain distribution of GABA receptors using the PET-FMZ (Fig. 1.A)^27^. We found a significant reduction (t(56)=8.83, P < 0.0001, Cohen d=2.32) of GABA_A_ receptor binding in the right (lesioned) hemisphere (Fig. 1.B). Importantly, TMS caused a reduction in GABA receptor binding variance (Fig. 1.C).

We then investigated if the changes in neurotransmission, suggesting synaptic adjustment, were associated with changes in brain activity consistent with the imbalance hypothesis. Specifically, we expected that TMS inhibitory stimulation to the healthy hemisphere should restore balance, because of disinhibition of the contralateral lesioned hemisphere, leading to rebalance ^33^. We found that the target (healthy) hemisphere was significantly activated during the saccade task in both TMS conditions (GLM t-test: 8>t(12)> 3.20; P<0.008, d=0.46) and that a task and TMS condition interaction was observed, corroborating an effect of TMS in the left (healthy) IPS (Fig. 2. A). Importantly, we found that activity levels increased 6-fold (compared to no change in the healthy hemisphere), for Real ipsilateral inhibitory stimulation in the contralateral (lesioned) hemisphere, suggesting contralateral disinhibition. This was confirmed by a random effects general linear model analysis of the fMRI data (see neural activation maps for both conditions, TMS sham and real; Fig. 2 B. and C.). This indicates a significant activation for the right (lesion) hemisphere specifically for the TMS real condition. The time-course activations show a striking stimulation dependence of activity in the contralateral right hemisphere. Accordingly, the right (lesioned) hemisphere is significantly activated only after real inhibitory TMS (t(12)>3.20, P<0.008, d=2; Fig. 2. C), in line with the interhemispheric disinhibition hypothesis.

**Figure. 2.**
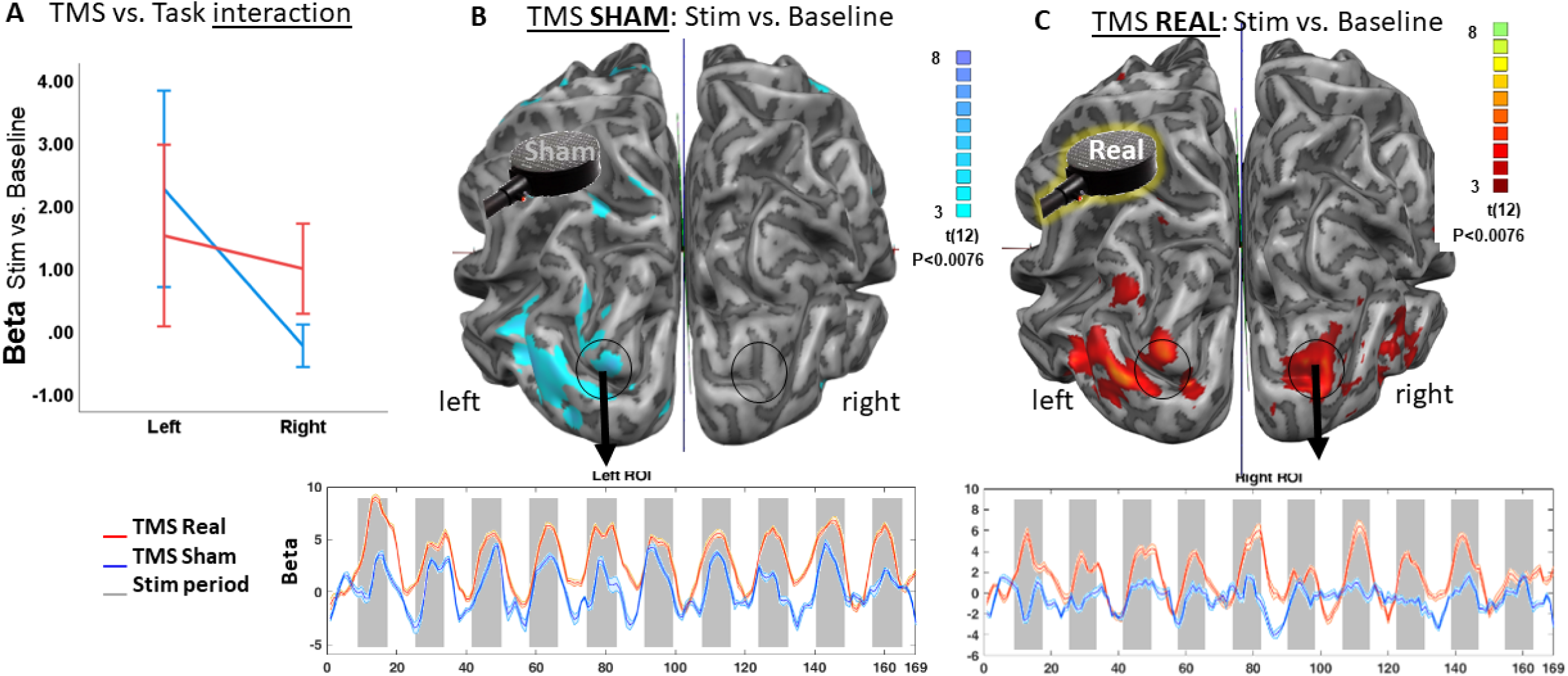
TMS rebalances brain activation during a Saccadic task. (A) Activation changes during the eye movement task: we found a significant interaction effect of the saccade task and TMS (F(1,11)=10, p<0.008, d=0.46), corroborating the direct impact of TMS. (B) TMS sham: brain activation during the saccade task (stimuli > baseline) was increased in the oculomotor network as expected, including frontal eye fields, for the target (left) hemisphere (where there is no lesion). No activation was found in the right hemisphere, possibly due to full inhibitory control of the healthy hemisphere during the task. The activation time-course for a left Region of Interest (ROI) is shown in the bottom plot: the grey bars represent the time periods where activation should increase. Note the similar activation patterns between TMS conditions. (C) TMS real: brain activation during the saccade task (stimuli > baseline) was increased in both hemispheres. Activation was also found in the right (lesioned) hemisphere. The activation time-course reveals a significant effect of the stimulation for the right ROI, as depicted in the plot. This map reveals a positive effect of the stimulation (inhibition of the target hemisphere) leading to a positive impact in the lesioned (right) hemisphere. Results are shown at P<0.008 corrected at cluster level (cluster defining threshold P<0.05). Error bars in the time-course plots reflect the SEM (standard error of the mean). Random effects (RFX) BOLD analysis.

The stimulation target region was the left IPS, and we performed region-of-interest (ROI)-based analyses (IPS left and right) of PET, and MRS GABA and Glx (Glutamatergic substances) data. ROIs were generated for PET analyses based on the MRS regions (Fig. 3.A). ROI analysis of the PET data did reveal significant differences in receptor level variance but not in their means between the TMS real and sham conditions (Fig. 3.B), while the MRS, allowed detecting consistent neurochemical changes following the TMS. Accordingly, a planned group analysis of Glx levels (Fig. 3. C) revealed a differential response between TMS real and TMS sham stimulation, with significantly reduced concentration of glutamatergic molecules (Glx) after real stimulation for both hemispheres (F(1,22)=8.90; P=0.009, d=0.26). Strikingly, TMS significantly reduced also the GABA variance in the right hemisphere (Levene F(1,14)=5.46, P=0.035, d=0.88; Fig. 3.D), showing that reduction of postsynaptic variance (as identified in PET data) is also accompanied by a similar effect at the presynaptic level. This was further confirmed by the reduction of Glx/GABA ratio (Left Sham: 4.67 ± 1.39 (Mean±Standard error); Left Real: 4.40±0.93; Right Sham: 4.30 ± 0.76; Right real: 3.53 ±0.22).

**Figure. 3.**
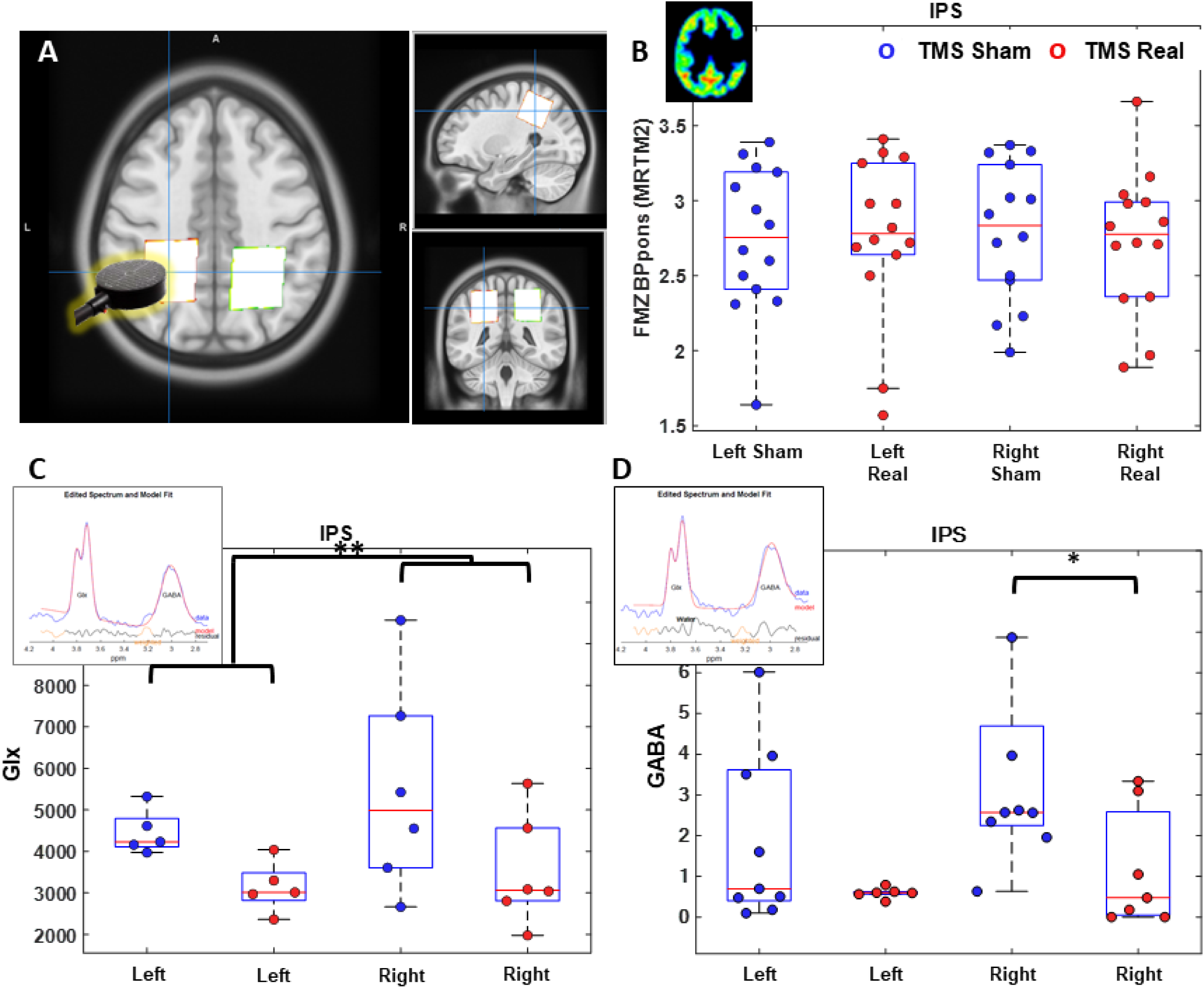
PET 11C-FMZ and MRS measures in IPS: impact of TMS on Glutamate levels and GABA neurotransmitter level variance. (**A**) Representation of the IPS regions in a standard MNI brain, used in the PET and MRS quantifications. (**B**) PET-FMZ data did not show significant differences between left/right IPS neither TMS real vs. sham in this region. (**C**) Glx (glutamatergic species) study and example spectrum: we found reduced Glx with TMS (Glx TMS sham vs. real (Left AND Right): F(1,22)=8.90; P=0.009; effect size d= 0.262). MRS data were acquired with the HERMES sequence: the fit output for the Glx and GABA signals are shown on the plots. Spectra were processed using the Gannet 3.0 toolkit, the blue line shows the raw pre-processed data, the red line is the fit of the model, and the black line is the residual difference between the experimental data and the curve fit. (**D**) GABA quantification results. Data revealed a significant effect of the TMS protocol on GABA level variance in the right (lesioned) hemisphere (contralateral to the stimulation) (Levene F(1,14)=5.46, P=0.035, d=0.88).

The association between pre and postsynaptic effects was confirmed by the finding that GABA levels were associated with GABA-A receptor binding only when TMS is real (r(11)=0.6424, p(Spearman)=0.037). This effect mainly stems from the lesioned hemisphere when TMS is real (r(6)=0.518, p(Spearman)=0.048) suggesting a long-range effect of TMS on regulation of neurotransmission (Fig. 4).

**Fig. 4.**
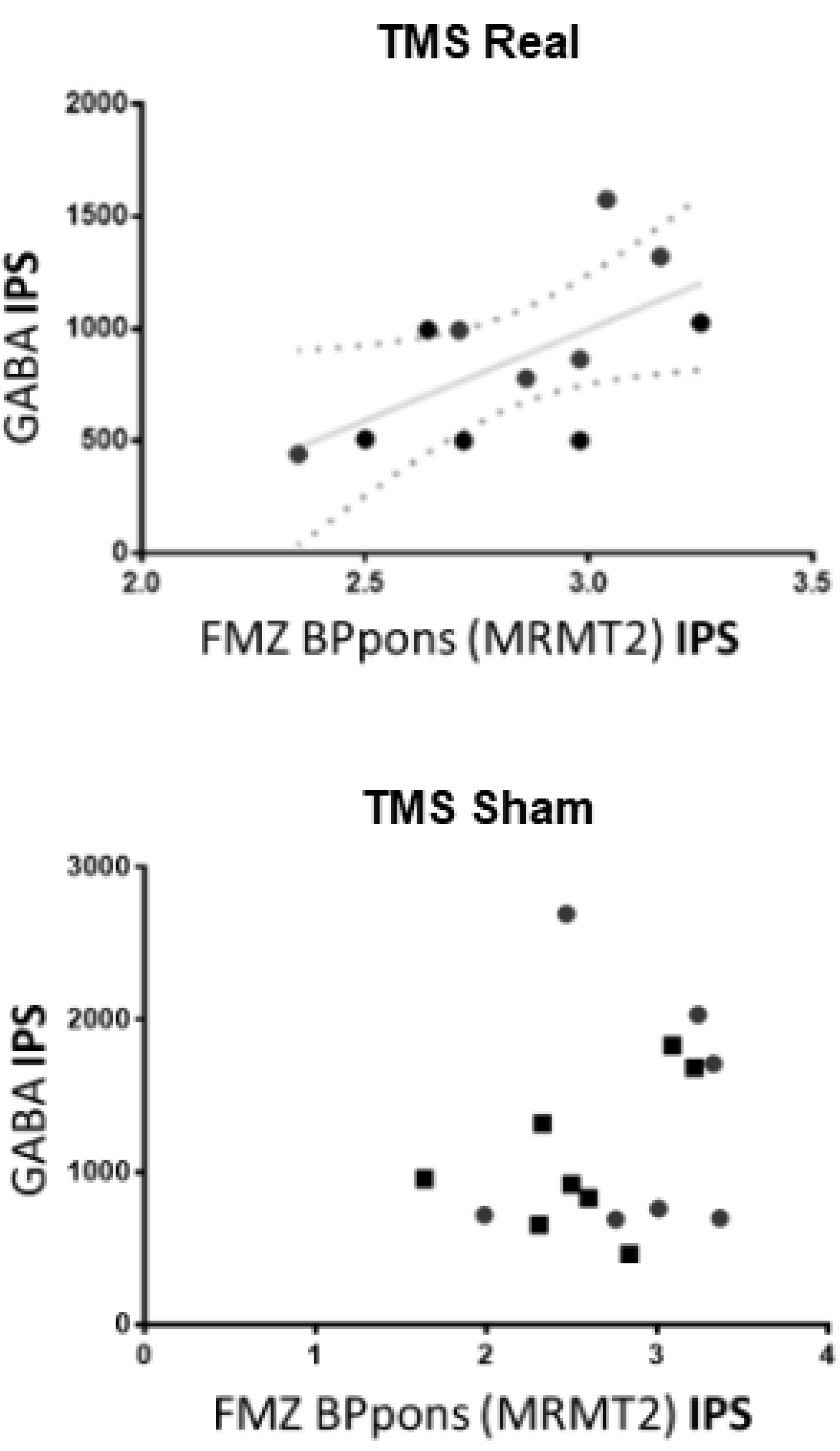
Correlations of PET and MRS data showed association of pre and postsynaptic measures suggestive of increased covariance across synaptic compartments. Scatter plots of PET-FMZ (GABAa receptors) vs. MRS (GABA) quantification in the IPS region of interest are shown for both TMS conditions. Significant correlation of GABA receptor binding (PET) and GABA (MRS) levels is present only when TMS is Real (r(11)=0.64, P (Spearman)=0.037).

In sum, real stimulation led to increased coupling between GABA neurotransmitter levels and its receptors, suggesting fine tuning of inhibitory neurotransmission. This aligns with the hypothesis of modulation of the (im)balance between hemispheres, as predicted by the inter-hemispheric inhibition model, supporting the use of TMS as a potential treatment approach to disorders of hemispheric asymmetry. This modulation of hemispheric interactions was associated with synaptic sharpening, a key mechanism associated with neuroplastic stabilization^19^.

The functional significance of GABA receptor levels was established at baseline by the observation that they were significantly associated with neuropsychological scores of parietal (left IPS) function in the absence of TMS (Landmark test, r(8)=0.81, P=0.015) and right parietal (IPS; Token test total score v r(7)=-0.777, P=0.04).

## Discussion

Our multimodal molecular imaging, spectroscopic and functional imaging data from the same day in stroke patients submitted to sham or real stimulation provide evidence for pre and postsynaptic sharpening at inhibitory synapses which is a mechanism of neuroplasticity ^19^ consistent with a model of a statistically optimal excitation/inhibition balance through variance reduction ^8^. Our results provide a mechanism of action which has direct implications for treatment approaches in stroke relying on interhemispheric inhibition using transcranial magnetic stimulation (TMS).

Our study generalizes to humans the notion that relevant synaptic parameters include variance, and covariance in neural measures ^4^ which are relevant for synaptic plasticity ^5^. Our TMS protocol likely modulated the number of synaptic inputs for which prior modelling work suggested that they depend on variability of postsynaptic responses ^6^. We hypothesize that decreased variability of drive to interneurons in the non-lesioned hemisphere, may help shape short and long-term plasticity. Our results support the interpretation of a recent postmortem and computational study in schizophrenia which showed that increased variability of excitatory drive to fast-spiking parvalbumin interneurons, leads to lower prefrontal gamma power^7^ which is known to be important to promote plasticity^34^.

Optimization of Synaptic coupling across hemispheres would favor plasticity through a statistically optimal E/I balance. Variability of pre/post changes due to optimizing postsynaptic response statistics (including enhanced co-variance, as shown by our data) leads to a framework where the full distribution of postsynaptic responses (instead of merely the mean weight) is optimized through joint pre- and postsynaptic modifications that are governed by a set of tightly coordinated neurotransmitters.

Furthermore, we found functional evidence for the interhemispheric inhibition model and a mechanism of action for inhibitory TMS in the hemisphere contralateral to the lesion in stroke. The bilateral response to a unilateral TMS can be interpreted in several ways. We show that inhibitory TMS induced improved coupling between GABAergic pre and postsynaptic function, and enhanced contralateral activation, thereby restoring hemispheric balance in association with reduction of molecular signal to noise ratios. Moreover, we demonstrate that our protocol was truly inhibitory, showing reduction in the excitatory glutamate/glutamine pool within the target (healthy) region, and recovery of neural responses in the contralateral (lesioned) regions in unilateral stroke. This empirically confirms the model of interhemispheric inhibition in vivo, coupled with pre and postsynaptic tuning. Our data further reveal that non-invasive brain stimulation is able to recover hemispheric balance upon unilateral lesions, even though our study being limited by the fact that precise localization of the site of stimulation (e.g. using MRI scan) was not performed. We tried to overcome this using a specific round coil, to ensure that we have stimulated relevant parts of parietal cortex in all patients. A more focal approach using a figure-of-eight coil would have run the risk of missing the target. Moreover, bilateral response to a unilateral TMS stimulation can vary depending on the specific location of the stimulation, the intensity of the stimulation and the characteristics of the individual being studied and thus this can be a limitation of our study.

A striking aspect of our data is the relatively long-lasting duration of neuronal and hemodynamic effects induced by a short TMS protocol: we applied TMS few minutes before each PET scan and approximately 2 hours before MRI (with significant effects in the lesioned hemisphere). This extends the timeline suggested by studies using similar stimulation protocols ^35,36^.

This ability of TMS to alter neurotransmission as demonstrated here suggests that transcranial brain stimulation can indeed be used to foster brain plasticity ^36^ in a number of neurological and neuropsychiatric disorders ^33,37^. Variance reduction has indeed been shown to represent a robust neuroplasticity mechanism *4*, *5, 6*. Uncovering determinants of these effects can only be achieved using a multimodal imaging approach ^39^ ^40^ ^41^.

As for the principle of brain lateralization, using an innovative multimodal approach in a unique patient group, our data provides direct evidence in favor of the interhemispheric inhibition model of hemispheric asymmetry, and its association with synaptic plasticity mechanisms, paving the way for mechanism driven clinical applications to treat disorders of interhemispheric imbalance, in particular in stroke.

## Data Availability

Data will be made available upon reasonable request

## Acknowledgements

We all the patients for their participation in the project.

## Funding

This project was supported by the grants from the European Commission, (EU H2020 AIMS-2-Trials and H2020-MSCA Marie Curie), FLAD Foundation (Life Sciences 2020) and Foundation for Science and Technology (FCT PAC MEDPERSYST 16428, DSAIPA/DS/0041/2020. FCT/UID4950 B&P. FCT 10.54499/CEECINST/00117/2021/CP2784/CT0001) and ICNAS-PHARMA.

## Competing interests

Authors declare that they have no competing interests.

## Supplementary material

Supplementary figures S1 and S2, and Table S1 are available at *Stroke* online.

## Notes

### Competing Interest Statement

The authors have declared no competing interest.

### Author Declarations

Comissão de Ética da Faculdade de Medicina da Universidade de Coimbra

## References

1. Karolis VR, Corbetta M, Thiebaut de Schotten M. The architecture of functional lateralisation and its relationship to callosal connectivity in the human brain. Nat Commun. 2019;10(1):1–9. doi:10.1038/s41467-019-09344-1

2. Güntürkün O, Ströckens F, Ocklenburg S. Brain Lateralization: A Comparative Perspective. Physiol Rev. 2020;100(3):1019–1063. doi:10.1152/PHYSREV.00006.2019

3. Ferro M, Lamanna J, Spadini S, Nespoli A, Sulpizio S, Malgaroli A. Synaptic plasticity mechanisms behind TMS efficacy: insights from its application to animal models. J Neural Transm. 2022;129(1):25–36. doi:10.1007/s00702-021-02436-7

4. Scheuss V, Neher E. Estimating Synaptic Parameters from Mean, Variance, and Covariance in Trains of Synaptic Responses. Biophys J. 2001;81(4):1970–1989. doi:10.1016/S0006-3495(01)75848-1

5. van Huijstee AN, Kessels HW. Variance analysis as a tool to predict the mechanism underlying synaptic plasticity. J Neurosci Methods. 2020;331:108526. doi:10.1016/j.jneumeth.2019.108526

6. Kretzberg J, Sejnowski T, Warzecha AK, Egelhaaf M. Variability of postsynaptic responses depends non-linearly on the number of synaptic inputs. Neurocomputing. 2003;52-54:313–320. doi:10.1016/S0925-2312(02)00797-X

7. Chung DW, Geramita MA, Lewis DA. Synaptic Variability and Cortical Gamma Oscillation Power in Schizophrenia. Am J Psychiatry. 2022;179(4):277–287. doi:10.1176/appi.ajp.2021.21080798

8. Costa RP, Padamsey Z, D’Amour JA, Emptage NJ, Froemke RC, Vogels TP. Synaptic Transmission Optimization Predicts Expression Loci of Long-Term Plasticity. Neuron. 2017;96(1):177–189.e7. doi:10.1016/j.neuron.2017.09.021

9. Malinow R, Tsien RW. Presynaptic enhancement shown by whole-cell recordings of long-term potentiation in hippocampal slices. Nature. 1990;346(6280):177-180. doi:10.1038/346177a0

10. Nowak DA, Grefkes C, Ameli M, Fink GR. Interhemispheric competition after stroke: Brain stimulation to enhance recovery of function of the affected hand. Neurorehabil Neural Repair. 2009;23(7):641–656. doi:10.1177/1545968309336661

11. Casula EP, Pellicciari MC, Bonnì S, et al. Evidence for interhemispheric imbalance in stroke patients as revealed by combining transcranial magnetic stimulation and electroencephalography. Hum Brain Mapp. 2021;42(5):1343–1358. doi:10.1002/hbm.25297

12. Xu J, Branscheidt M, Schambra H, et al. Rethinking interhemispheric imbalance as a target for stroke neurorehabilitation. Ann Neurol. 2019;85(4):502–513. doi:10.1002/ana.25452

13. Dionísio A, Gouveia R, Castelhano J, et al. The Role of Continuous Theta Burst TMS in the Neurorehabilitation of Subacute Stroke Patients: A Placebo-Controlled Study. Front Neurol. 2021;12. doi:10.3389/fneur.2021.749798

14. Dionísio A, Gouveia R, Duarte IC, Castelhano J, Duecker F, Castelo-Branco M. Continuous theta burst stimulation increases contralateral mu and beta rhythms with arm elevation: implications for neurorehabilitation. J Neural Transm. 2020;127(1):17–25. doi:10.1007/s00702-019-02117-6

15. Dionísio A, Gouveia R, Castelhano J, et al. The Neurophysiological Impact of Subacute Stroke: Changes in Cortical Oscillations Evoked by Bimanual Finger Movement. Stroke Res Treat. 2022;2022. doi:10.1155/2022/9772147

16. Vidal AC, Banca P, Pascoal AG, et al. Bilateral versus ipsilesional cortico-subcortical activity patterns in stroke show hemispheric dependence. Int J Stroke. 2017;12(1):71–83. doi:10.1177/1747493016672087

17. Vidal AC, Banca P, Pascoal AG, Cordeiro G, Sargento-Freitas J, Castelo-Branco M. Modulation of Cortical Interhemispheric Interactions by Motor Facilitation or Restraint. Neural Plast. 2014;2014:1–8. doi:10.1155/2014/210396

18. Dionísio A, Duarte IC, Patrício M, Castelo-Branco M. The Use of Repetitive Transcranial Magnetic Stimulation for Stroke Rehabilitation: A Systematic Review. J Stroke Cerebrovasc Dis. 2018;27(1):1–31. doi:10.1016/j.jstrokecerebrovasdis.2017.09.008

19. Dionísio A, Duarte IC, Patrício M, Castelo-Branco M. Transcranial Magnetic Stimulation as an Intervention Tool to Recover from Language, Swallowing and Attentional Deficits after Stroke: A Systematic Review. Cerebrovasc Dis. 2018;46(3-4):176–183. doi:10.1159/000494213

20. Pascual-Leone A, Walsh V, Rothwell J. Transcranial magnetic stimulation in cognitive neuroscience - Virtual lesion, chronometry, and functional connectivity. Curr Opin Neurobiol. 2000;10(2):232–237. doi:10.1016/S0959-4388(00)00081-7

21. Rossi S, Hallett M, Rossini PM, Pascual-Leone A. Safety, ethical considerations, and application guidelines for the use of transcranial magnetic stimulation in clinical practice and research. Clin Neurophysiol. 2009;120(12):2008–2039. doi:10.1016/j.clinph.2009.08.016

22. Di Gregorio F, La Porta F, Casanova E, et al. Efficacy of repetitive transcranial magnetic stimulation combined with visual scanning treatment on cognitive and behavioral symptoms of left hemispatial neglect in right hemispheric stroke patients: study protocol for a randomized controlled trial. Trials. 2021;22(1):24. doi:10.1186/s13063-020-04943-6

23. Sack AT, Linden DEJ. Combining transcranial magnetic stimulation and functional imaging in cognitive brain research: Possibilities and limitations. Brain Res Rev. 2003;43(1):41–56. doi:10.1016/S0165-0173(03)00191-7

24. Fried P, Jannati A, Morris T, et al. Relationship of active to resting motor threshold influences the aftereffects of theta-burst stimulation. Brain Stimul. 2019;12(2):465. doi:10.1016/J.BRS.2018.12.513

25. Cole EJ, Phillips AL, Bentzley BS, et al. Stanford Neuromodulation Therapy (SNT): A Double-Blind Randomized Controlled Trial. Am J Psychiatry. 2022;179(2):132–141. doi:10.1176/APPI.AJP.2021.20101429/ASSET/IMAGES/MEDIUM/APPI.AJP.2021.20101429F1.GIF

26. Blumberger DM, Vila-Rodriguez F, Thorpe KE, et al. Effectiveness of theta burst versus high-frequency repetitive transcranial magnetic stimulation in patients with depression (THREE-D): a randomised non-inferiority trial. Lancet. 2018;391(10131):1683–1692. doi:10.1016/S0140-6736(18)30295-2

27. Violante IR, Patricio M, Bernardino I, et al. GABA deficiency in NF1. Neurology. 2016;87(9):897–904. doi:10.1212/WNL.0000000000003044

28. Edden R a. E, Puts N a. J, Harris AD, Barker PB, Evans CJ. Gannet: A batch-processing tool for the quantitative analysis of gamma-aminobutyric acid-edited MR spectroscopy spectra. J Magn Reson Imaging. 2013;00:n/a-n/a. doi:10.1002/jmri.24478

29. Aufhaus E, Weber-Fahr W, Sack M, et al. Absence of changes in GABA concentrations with age and gender in the human anterior cingulate cortex: a MEGA-PRESS study with symmetric editing pulse frequencies for macromolecule suppression. Magn Reson Med. 2013;69(2):317–320. doi:10.1002/MRM.24257

30. Kutner MH, Nachtsheim CJ, Neter J, et al. Applied Linear Statistical Models Fifth Edition t:a Irwin. Published online 2005. Accessed December 13, 2021. www.mhhe.com

31. Friston KJ, Josephs O, Rees G, Turner R. Nonlinear event-related responses in fMRI. Magn Reson Med. 1998;39(1):41–52. doi:10.1002/MRM.1910390109

32. Lenoski B, Baxter LC, Karam LJ, Maisog J, Debbins J. On the performance of autocorrelation estimation algorithms for fMRI analysis. IEEE J Sel Top Signal Process. 2008;2(6):828–838. doi:10.1109/JSTSP.2008.2007819

33. Chistyakov a V, Soustiel JF, Hafner H, Trubnik M, Levy G, Feinsod M. Excitatory and inhibitory corticospinal responses to transcranial magnetic stimulation in patients with minor to moderate head injury. J Neurol Neurosurg Psychiatry. 2001;70(5):580–587. http://www.pubmedcentral.nih.gov/articlerender.fcgi?artid=1737339&tool=pmcentrez&rendertype=abstract

34. Traub RD, Spruston N, Soltesz I, Konnerth A, Whittington MA, Jefferys JG. Gamma-frequency oscillations: a neuronal population phenomenon, regulated by synaptic and intrinsic cellular processes, and inducing synaptic plasticity. Prog Neurobiol. 1998;55(6):563–575. doi:10.1016/S0301-0082(98)00020-3

35. Pascual-Leone A, Valls-Sole J, Wassermann EM, Hallett M. Responses to Rapid-Rate Transcranial Magnetic Stimulation of the Human Motor Cortex. Vol 117.; 1994. Accessed March 26, 2021. http://brain.oxfordjournals.org/

36. Huang YZ, Edwards MJ, Rounis E, Bhatia KP, Rothwell JC. Theta burst stimulation of the human motor cortex. Neuron. 2005;45(2):201–206. doi:10.1016/j.neuron.2004.12.033

37. McClintock SM, Freitas C, Oberman L, Lisanby SH, Pascual-Leone A. Transcranial magnetic stimulation: a neuroscientific probe of cortical function in schizophrenia. Biol Psychiatry. 2011;70(1):19–27. doi:10.1016/j.biopsych.2011.02.031

38. Parker D, Bevan S. Modulation of Cellular and Synaptic Variability in the Lamprey Spinal Cord. J Neurophysiol. 2007;97(1):44–56. doi:10.1152/jn.00717.2006

39. Rossini PM, Altamura C, Ferreri F, et al. Neuroimaging experimental studies on brain plasticity in recovery from stroke. Eura Medicophys. 2007;43(2):241–254. Accessed March 19, 2021. https://pubmed.ncbi.nlm.nih.gov/17589415/

40. Allen EA, Pasley BN, Duong T, Freeman RD. Transcranial magnetic stimulation elicits coupled neural and hemodynamic consequences. Science (80-). 2007;317(5846):1918–1921. doi:10.1126/science.1146426

41. Sack AT, Kohler A, Bestmann S, et al. Imaging the Brain Activity Changes Underlying Impaired Visuospatial Judgments: Simultaneous fMRI, TMS, and Behavioral Studies. Cereb Cortex. 2007;17(12):2841–2852. doi:10.1093/cercor/bhm013

